# Cognitive abilities and irritability are the main factors influencing initial placement of autistic preschoolers in special or mainstream education

**DOI:** 10.1101/2025.08.27.25334558

**Authors:** Moran Bachrach, Michal Ilan, Michal Faroy, Analya Michaelovsky, Dikla Zagdon, Yair Sadaka, Omer Bar Yosef, Adi Aran, Michal Begin, Ditza Zachor, Einat Avni, Judah Koller, Idan Menashe, Gal Meiri, Ilan Dinstein

## Abstract

In many countries, autistic children are placed in either exclusive special education or inclusive mainstream preschool settings following diagnosis. These settings differ in their staff composition and expertise, ability to implement structured autism interventions, ability to integrate autistic and typically developing children, and costs. Here, we examined whether there were significant differences in the behavioral abilities and developmental difficulties of children placed in either setting in Israel. We analyzed data from 165 autistic children, 120 in special and 45 in mainstream education, who completed comprehensive behavioral assessments at a mean age of 37.8 months, as they entered their first preschool setting. Children placed in special education exhibited significantly poorer cognitive abilities and higher irritability and hyperactivity than children in mainstream education while there were no significant differences in autism severity or adaptive behaviors across groups. Moreover, cognitive and irritability scores were sufficient for classifying children across the two settings when using a pruned decision tree algorithm and a 5-fold cross-validation procedure. These findings extend previous research by demonstrating that cognitive abilities and irritability are the strongest predictors of initial educational placement. Further longitudinal research is needed to determine whether these placement decisions benefit the children as they develop.

## Introduction

Children diagnosed with autism are remarkably heterogeneous, exhibiting varying levels of social difficulties, restricted and repetitive behaviors (RRBs), cognitive abilities, language skills, adaptive behaviors, and aberrant behaviors (Trembath & Vivanti, 2014; Zachor & Ben-Itzchak, 2017). In many countries preschool children with autism are placed in either inclusive mainstream education or exclusive special education settings (Van Kessel et al., 2019). There are currently no clear guidelines or criteria for placing preschool children with specific characteristics in either setting (Rattaz et al., 2020; Towle et al., 2018; White et al., 2007).

In Israel and many other countries, there are dramatic differences between government-funded special and mainstream educational settings. Special education offers an exclusive environment where a small number of autistic children (typically ≤8) receive structured autism-specific interventions from a highly professional multidisciplinary educational staff who tailor the intervention to the specific needs of each child (Arnold et al., 2021). Many studies have demonstrated that early autism interventions improve later outcomes (Gabbay-Dizdar et al., 2022; Hyman et al., 2020; Zwaigenbaum et al., 2015), thereby motivating the investment in specialized autism education settings. In Israel, approximately two third of autistic children are placed in such specialized setting after receiving an autism diagnosis (Ilan et al., 2021).

In contrast, inclusive mainstream preschools accommodate larger groups of children with fewer staff who are usually not trained to provide autism-specific intervention. Autistic children who are integrated in these settings often qualify for a personal aid to assist with daily activities and facilitate peer interactions (Barton et al., 2012; Lynch & Irvine, 2009), but teachers often report that integration is challenging (Gavaldá & Qinyi, 2012; Lindsay et al., 2014). Despite these challenges, autistic children in integrated settings can benefit from exposure to their typically developing peers in many ways (Arnold et al., 2021; Farrell, 2000; Sansour & Bernhard, 2018). Note that mainstream education settings are considerably cheaper to create and maintain. It has been estimated that placing a child with autism in special education costs 4 times more than placement in mainstream education (Chasson et al., 2007).

Relatively few studies have examined how children’s characteristics are associated with educational placement and previous studies have mostly focused on older school-aged children rather than preschool children. Studies from France and the U.S. have reported that school-aged children with lower cognitive abilities are more likely to be placed in special education rather than mainstream settings (Rattaz et al., 2020; White et al., 2007), while studies in Denmark (Christiansen et al., 2021) and Canada (Lyons et al., 2011) reported that more severe autism symptoms and poorer social competence were associated with placement in special education schools. In a previous study, we reported that Israeli preschool autistic children placed in special education had significantly lower cognitive abilities than those placed in mainstream education settings, but did not differ in the severity of core autism symptoms (Ilan et al., 2021). To the best of our knowledge, previous studies have not examined the potential impact of adaptive or aberrant behaviors on placement in preschool settings.

Adaptive behavior difficulties are common in autistic preschoolers and often include poor daily living skills, communication, and socialization skills (Stone et al., 1999). Challenges in these domains are associated with autism severity, low social interest, and behavioral problems (Franchini et al., 2018), and suggest that children with poor adaptive abilities may require more support in their educational settings. However, while some studies have reported that school-aged autistic children with poorer adaptive skills were more likely to be placed in special education settings (Towle et al., 2018; White et al., 2007), others did not (Rattaz et al., 2020).

Aberrant behaviors are also common in autistic children (Kozlowski & Matson, 2012) and include aggressive, confrontational, and self-injurious behaviors as well as hyperactivity (Bauminger et al., 2010; Emerson et al., 2001; Kaat & Lecavalier, 2013). In older school-aged children, these behaviors hinder academic engagement and prosocial interactions and create formidable barriers to inclusion in mainstream educational settings (Boyd et al., 2012; Dunlap et al., 2010; Sterling-Turner et al., 2001), sometimes motivating intervention with pharmacological treatments (Haem et al., 2020; Kaat & Lecavalier, 2013). However, other studies have not found differences in aberrant behaviors across school-aged children placed in special versus mainstream educational settings (Rattaz et al., 2020).

Given the limited literature on factors that influence early preschool placement, we systematically examined whether cognitive abilities, core autism symptoms, adaptive behaviors, and aberrant behaviors differed across a relatively large sample of 3-year-old autistic children entering their first year of preschool in either a special or mainstream education setting.

## Methods

### Participants and procedures

We recruited 165 children following their autism diagnosis as they entered their first year of preschool education at an average age of 37.86 (SD=4.4) months. Of these children, 120 (73%) were placed in special education and 45 (27%) were placed in mainstream education settings. Recruitment was performed by the Azrieli National Centre for Autism and Neurodevelopment Research (ANCAN), an ongoing collaboration between Ben-Gurion University of the Negev (BGU) and nine clinical sites throughout Israel where autistic children are diagnosed and treated (Dinstein et al., 2020). ANCAN regularly recruits children for a variety of research projects and manages the National Autism Database of Israel (NADI), which holds anonymized retrospective information from thousands of autistic children and their family members (Meiri et al., 2017). All children aged 27-47 months, who were diagnosed at ANCAN partner sites between 2017 and 2024, completed an Autism Diagnostic Observation Schedule, 2^nd^ edition (ADOS-2) assessment, and the child’s initial educational placement was known/recorded, were included in the study. All children had a formal diagnosis of autism as established by both a developmental psychologist and a pediatric neurologist or psychiatrist. All were born full term without major complications (i.e., 36-42 weeks gestation age and birth weight >2500 grams) and no known metabolic, neurological, or genetic syndromes. Of the selected 165 children, 114 (69%) completed cognitive assessments, 113 (68.5%) completed the Adaptive Behavior Rating Scale, 3^rd^ edition (ABAS-3), and 127 (77%) completed the Aberrant Behaviors Checklist (ABC). The Soroka University Medical Center (SUMC) Helsinki Committee approved this study.

### Measures

#### ADOS-2

Children were evaluated by an experienced clinician with research reliability using the toddler module or modules 1–3 of the ADOS-2 (Lord et al., 2012) according to their age and language abilities. We transformed raw ADOS-2 scores into Calibrated Severity Scores (CSS), which allow comparison of autism severity across children of different ages and language capabilities (Esler et al., 2015; Gotham et al., 2009). The ADOS-2 CSS was computed separately for social affect (SA) and restricted and repetitive behaviors (RRB) domains (Esler et al., 2015; Hus et al., 2014).

#### Cognitive assessments

Cognitive ability was measured using multiple tests depending on chronological and mental age, as determined by the developmental psychologist. The Bayley Scales of Infant and Toddler Development, Third Edition (Viezel et al., 2014) was used with 29 (∼25.4%) children, the Mullen Scales of Early Learning (MSEL;Mullen, 1995) was used with 65 (∼57%) children, and the Wechsler Preschool and Primary Scale of Intelligence, Third Edition(Luiselli et al., 2013) was used with 20 (∼17.6%) children. The three tests yield equivalent standardized scores with a mean of 100 and a standard deviation of 15. Since strong correlations exist between the Bayley and Wechsler tests, as well as between the Mullen and Bayley tests (Bayley, 2006; Lense et al., 2014), we combined scores from these tests in our analysis.

#### Adaptive behaviors

Adaptive behaviors were measured using the Adaptive Behavior Rating Scale, third edition (ABAS-3)(Balboni et al., 2014; Kane & Oakland, 2015). Parents completed the Parent/Primary Caretaker Form (0–5 years old), which includes 241 items and yields scores in 10 subscales (Communication, Community Use, Functional Pre-academics, Home Living, Health and Safety, Leisure, Self-Care, Self-Direction, Social and Motor), three composite score domains (Conceptual, Social, and Practical), and an overall General Adaptive Composite (GAC) score.

#### Aberrant behaviors

Aberrant behaviors were measured using the Aberrant Behaviors Checklist (ABC) (Brinkley et al., 2007; Rojahn et al., 2013) which was filled out by the parents. The ABC is a 58-question parent-behavior rating scale that yields scores in five subscales: (1) irritability; (2) social withdrawal; (3) stereotypical behavior; (4) hyperactivity; and (5) inappropriate speech (Aman, 1985).

#### Parental questionnaires

Parents completed questionnaires regarding parental age at the time of their child’s birth, parental education levels, socioeconomic status, average household income, and the educational placement setting their child had entered after diagnosis.

### Data analysis and statistics

All statistical analyses were conducted using RStudio (RStudio Inc., Boston, MA). We imputed missing data using the Random Forest algorithm (Stekhoven & Bühlmann, 2012) with 20 iterations. This approach generated a complete dataset by estimating plausible values for missing entries, given the statistics of existing data. Independent two-tailed t-tests (assuming unequal variances) were conducted to compare children placed in special and mainstream education settings (Table 1). We did not correct for multiple comparisons in these initial exploratory analyses to retain high sensitivity.

**Table 1:** Behavioral and Socio-Demographic Characteristics of children with autism across educational settings.

|  | Mainstream<br>(n=45,<br>males=34) | Special<br>(n=120,<br>males=96) | Statistics | *P* | Cohen's D |
| --- | --- | --- | --- | --- | --- |
| Measures | Mean (SD) |  |  |  |  |
| Assessments age (M) | 37.9 (5.13) | 37.8 (4.25) |  |  |  |
| **Age at diagnosis** | **33.5 (7.60)** | **30.2 (9.54)** | **t (98.69) = 2.29** | **0.024\*** | **0.36** |
| ADOS-2 CSS | 6.58 (2.03) | 7.03 (1.90) | t (74.83) = -1.28 | 0.20 | -0.23 |
| ADOS-2 SA CSS | 6.11 (2.01) | 6.45 (2.16) | t (84.24) = -0.94 | 0.34 | -0.15 |
| ADOS-2 RRB CSS | 7.69 (1.78) | 8.21 (1.48) | t (68.14) = -1.74 | 0.08 | -0.33 |
| **Cognitive score** | **79.9 (22.6)** | **67.8 (18.6)** | **t (67.67) = 3.18** | **0.002\*** | **0.60** |
| ABAS general score | 71.7 (18.5) | 66.6 (15.3) | t (67.95) = 1.66 | 0.10 | 0.31 |
| ABAS conceptual score | 75.2 (18.9) | 69.3 (15.1) | t (66.36) = 1.90 | 0.06 | 0.36 |
| ABAS social score | 72.6 (16.9) | 68.2 (13.7) | t (66.93) = 1.59 | 0.11 | 0.30 |
| ABAS practical score | 72.7 (17.2) | 68.1 (15.1) | t (70.89) = 1.59 | 0.11 | 0.29 |
| **ABC: Irritability score** | **9.00 (10.1)** | **13.6 (10.4)** | **t (81.45) = -2.58** | **0.011\*** | **-0.44** |
| ABC: Social withdrawal | 10.2 (12.9) | 11.0 (9.46) | t (62.57) = -0.38 | 0.70 | -0.07 |
| ABC: Stereotypic behavior | 4.80 (6.07) | 5.61 (5.41) | t (71.72) = -0.78 | 0.43 | -0.14 |
| **ABC: Hyperactivity /**<br>**Noncompliance** | **13.8 (11.1)** | **18.4 (11.9)** | **t (84.27) = -2.30** | **0.023\*** | **-0.39** |
| ABC: Inappropriate speech | 2.42 (3.14) | 3.20 (3.15) | t (79.09) = -1.41 | 0.16 | -0.24 |
| Socioeconomic level | 4.64(0.95) | 4.83 (1.15) | t (94.09) = -1.06 | 0.28 | -0.17 |

|  | Mainstream<br>(n=45,<br>males=34) | Special<br>(n=120,<br>males=96) | Statistics | *P* | Cohen's *D* |
| --- | --- | --- | --- | --- | --- |
| Measures | Mean (SD) |  |  |  |  |
| Average income | 1.51 (1.14) | 1.38 (0.96) | *t* (68.85) =0.66 | 0.50 | 0.12 |
| **Mother's years of education** | **14.5 (2.84)** | **13.6 (2.56)** | ***t* (72.45) = 1.98** | **0.050\*** | **0.36** |
| Father's years of education | 13.4 (3.13) | 13.1 (2.17) | *t* (60.49) = 0.69 | 0.48 | 0.14 |
| Mother's age at birth | 31.0 (5.44) | 32.3 (5.66) | *t* (81.93) = -1.36 | 0.17 | -0.23 |
| Father's age at birth | 33.6 (5.37) | 34.5 (5.94) | *t* (86.88) = -0.91 | 0.36 | -0.15 |

ADOS: Autism Diagnostic Observation Schedule; CSS: calibrated severity scores; SA: social affect; RRB: restricted and repetitive behaviors; ABAS: Adaptive Behavior Rating Scale; ABC: Aberrant Behaviors Checklist.

Next, we performed a classification tree analysis using the “rpart” package in R (Therneau & Atkinson, 2025) to classify children across educational settings. The classification model was trained on data from 15 measures: cognitive ability, ADOS-2 CSS, Social Affect and RRB scores, adaptive behavior indices (ABAS subscales: General score, Conceptual score, Social score and Practical score), aberrant behavior subscales (ABC subscales: Irritability score, Social withdrawal score, Stereotypic behavior score, Hyperactivity / Noncompliance score and Inappropriate speech score), and demographic variables: age at diagnosis and maternal education. Prior to training, we addressed class imbalance using the Synthetic Minority Over-sampling Technique (SMOTE) (Chawla et al., 2002). This method artificially increased the size of the underrepresented group (mainstream placement) by generating synthetic cases based on nearest neighbors, thereby balancing the two groups and improving classification robustness. A classification tree was then trained using recursive partitioning with the Gini impurity index as the splitting criterion (Greenwell, 2022). The model was trained and tested using a 5-fold cross-validation strategy where 80% of the children were used to train the model and classification accuracy was tested with the remaining 20% of children who were left out. Different training and testing samples were used in each of the 5 iterations and we report the average classification accuracy across iterations. We also computed an optimal complexity parameter (cp) for identifying a pruned classification tree with minimal overfitting to specific iterations. We computed the confusion matrix when analyzing the entire data with this optimized model.

## Results

In initial exploratory analyses, we compared behavioral and developmental scores between children in special versus mainstream educational settings using independent-samples *t*-tests (Figures 1&2). Children in special education had significantly lower cognitive scores (*t*(67.67) = 3.18, *p* = .002, *d* = 0.60), higher levels of irritability (*t*(81.45) = –2.58, *p* = .011, *d* = –0.44) and greater hyperactivity/noncompliance (*t*(84.27) = –2.30, *p* = .023, *d* = –0.39) compared to those in mainstream settings. In addition, children in special education were diagnosed at a significantly younger age (*t*(98.69) = 2.29, *p* = .024, *d* = 0.36) and their mothers had significantly less education (*t*(72.45) = 1.98, *p* = .050, *d* = 0.36). Note that we did not correct for multiple comparisons to retain high sensitivity.

**Figure 1:**
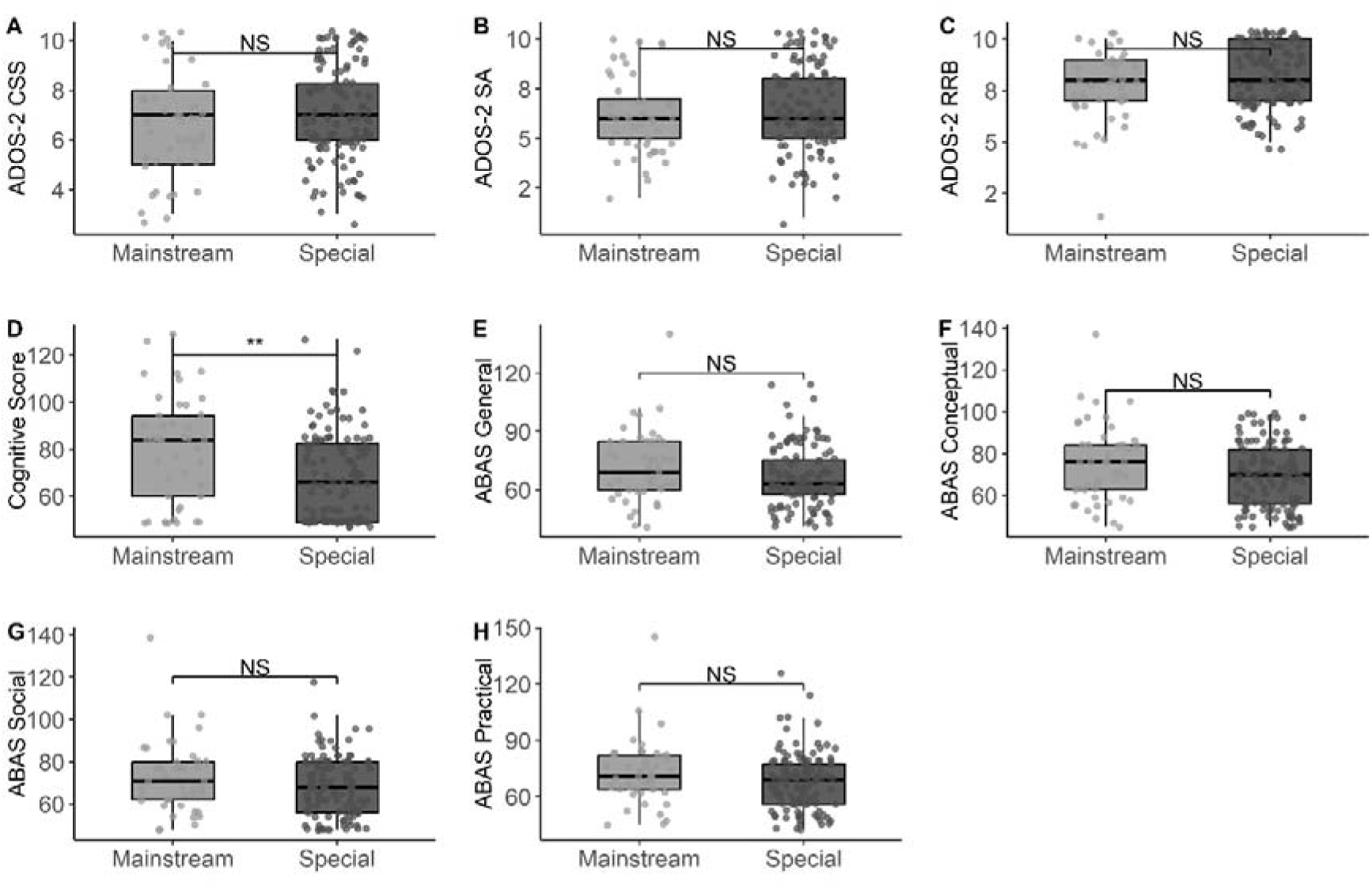
Comparison across children placed in mainstream and special education settings. Box and whisker plots demonstrate differences in (A) ADOS-2 CSS, (B) ADOS-2 SA-CSS, (C) ADOS-2 RRB-CSS, (D) Cognitive, (E) ABAS general, (F) ABAS conceptual, (G) ABAS social, and (H) ABAS practical scores.

**Figure 2:**
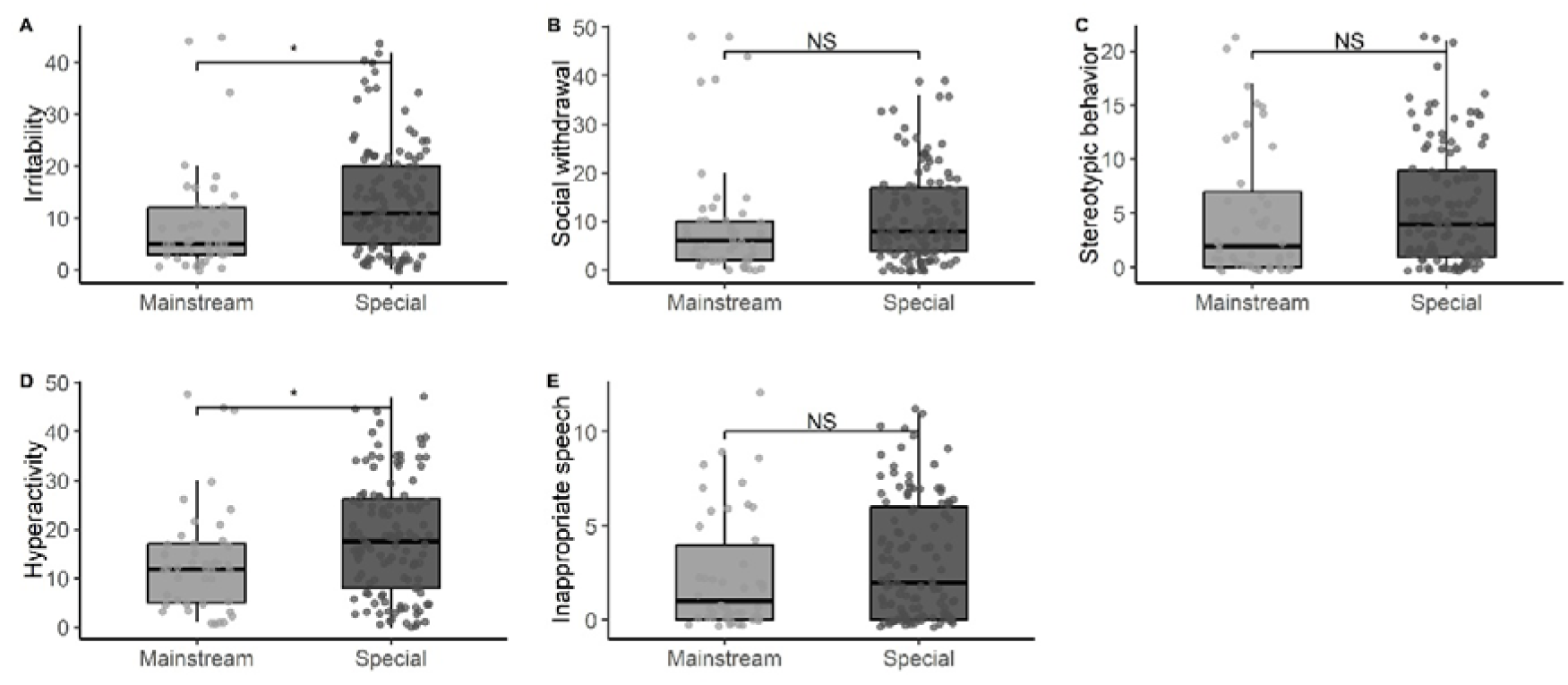
Differences in ABC scores across children placed in mainstream and special education settings. Box and whisker plots demonstrate differences in ABC subscale scores: (A) Irritability, (B) Social withdrawal, (C) Stereotypical behaviors, (D) Hyperactivity/ Noncompliance, and (E) Inappropriate speech.

In contrast, there were no significant group differences in total ADOS-2 CSS (*p* = .20), ADOS-2 SA CSS (*p* = .34), ADOS-2 RRB CSS (*p* = .08), ABAS general composite (*p* = .10), ABAS social (*p* = .11), ABAS practical (*p* = .11), ABAS conceptual (*p* = .06), ABC social withdrawal (*p* = .70), ABC stereotypic behavior (*p* = .43), or ABC inappropriate speech (*p* = .16) scores.

### Classifying initial preschool placement

Next, we used a decision tree analysis while applying a 5-fold cross-validation procedure to create a reliable classifier of educational placement (mainstream vs. special education). The classification model was trained with 15 variables, which included cognitive, ADOS-2, ABAS, and ABC scores as well as demographic characteristics including age of diagnosis and maternal education (see Methods). We balanced the data across the two classes (i.e., educational settings) using the Synthetic Minority Over-sampling Technique (SMOTE; Chawla et al., 2002) prior to training and testing.

The pruned classification tree model achieved an average classification accuracy of 76.4% (SD = 4.5%) and demonstrated fair agreement between predicted and actual placement (Kappa = 0.27). The final model first split the data according to cognitive ability, with all children who had cognitive scores < 84 being assigned to the special education group. Children with cognitive scores ≥ 84 were further split according to irritability scores, with all children who had irritability scores ≥ 8 assigned to the special education group (Figure 3A). The resulting confusion matrix (Figure 3B) yielded a sensitivity of 0.825 and specificity of 0.66 for correctly identifying children in special education with a positive predictive value of 0.87 and a negative predictive value of 0.73.

**Figure 3:**
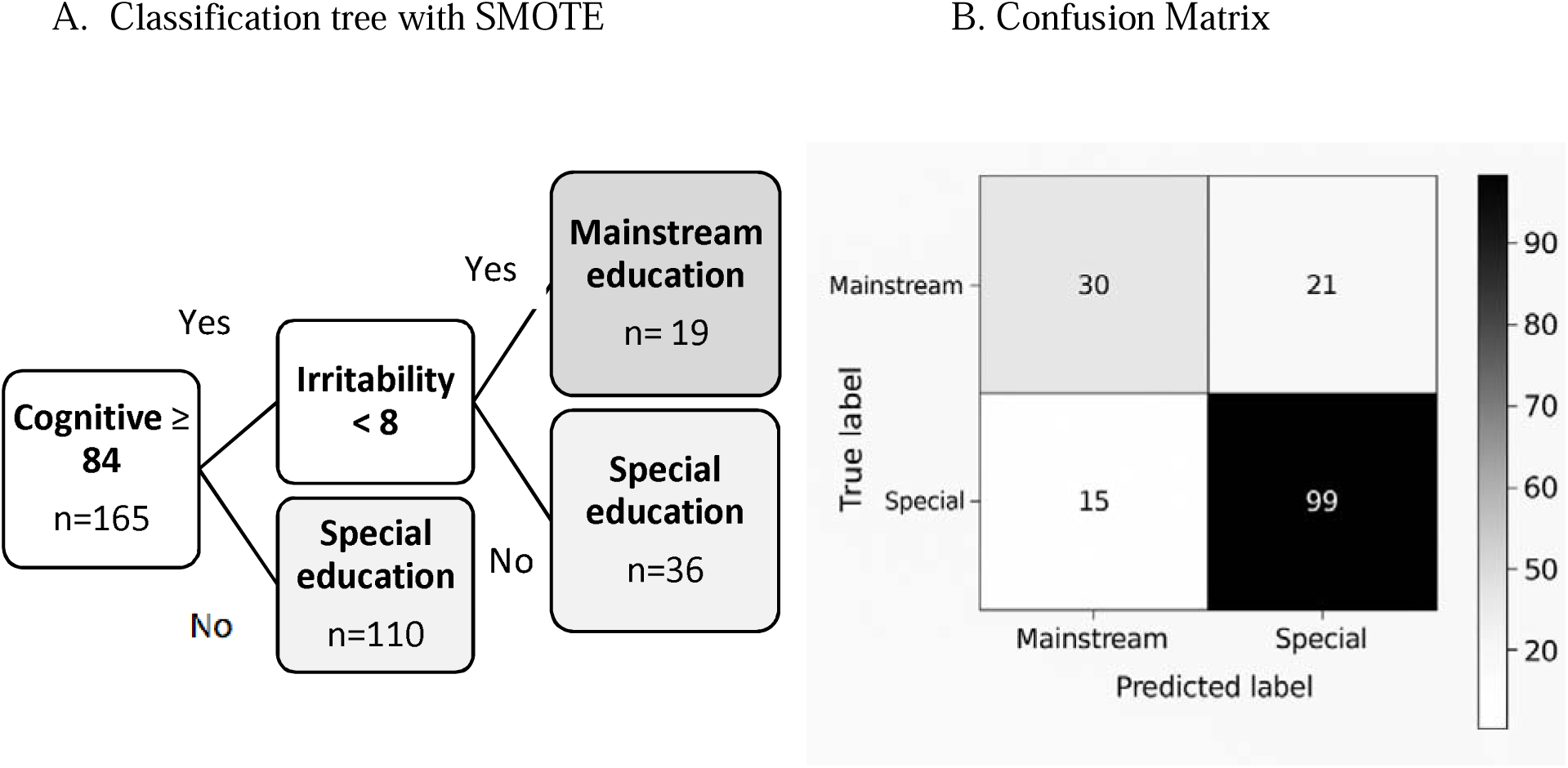
**A.** A pruned classification tree derived from 5-fold cross-validated analysis, predicting preschool educational placement (mainstream vs. special education) The model was trained on a balanced dataset and optimized using the complexity parameter that maximized cross-validated accuracy. **B.** Confusion matrix demonstrating the accuracy of the pruned decision tree model in predicting preschool educational placement based on cognitive scores and irritability alone.

## Discussion

Our results suggest that poor cognitive abilities and higher irritability are the main factors influencing the placement of preschool autistic children in special education, rather than autism severity or adaptive behaviors. This was apparent in exploratory univariate analyses (Figures 1&2) and confirmed by a multivariate classification analysis (Figure 3) using extensive data collected from 165 autistic children as they entered their first year in a preschool setting.

Previous studies with school-aged autistic children have reported mixed results regarding potential differences in cognitive abilities across children placed in special and mainstream education. While some have reported that cognitive scores were significantly lower in children placed in special education (Christiansen et al., 2021; Kim et al., 2018; Rattaz et al., 2020; White et al., 2007), others did not (Christiansen et al., 2021; Kurth & Mastergeorge, 2010). The only previous study examining preschool autistic children was from our lab and reported significantly poorer cognitive scores in children placed in special education (Ilan et al., 2021). Note that the current study replicates our previous findings with an independent sample of different children. This suggests that cognitive scores are indeed the strongest predictor of educational placement for preschoolers with autism.

Similarly, previous studies with school-aged children have reported mixed results regarding potential differences in autism severity across children placed in special and mainstream education. While some studies with school age children have reported that higher autism severity was associated with placement in special education (Christiansen et al., 2021; Lyons et al., 2011) others have not (Rattaz et al., 2020; White et al., 2007). Our previous study with preschool autistic children reported that ADOS-2 CSS did not differ significantly across children in special and mainstream educational settings (Ilan et al., 2021) and the current results replicate this finding with an independent sample. This suggests that autism severity is currently not a major consideration when selecting the initial preschool setting for autistic children.

An important contribution of the current study was the concomitant examination of aberrant behaviors and adaptive behaviors as potential factors that influence placement in special or mainstream education, which have not been examined previously in preschool children.

Previous studies with school aged children have reported that aberrant behaviors including aggression, self-injurious behavior, and emotional volatility disrupt classroom learning and social engagement (Brotman et al., 2017; Farmer & Aman, 2020) and exclude autistic children from mainstream education (Boyd et al., 2012; Dunlap et al., 2010). Our findings demonstrate that higher irritability, in particular, is associated with placement of autistic preschoolers in special education. Note that other ABC subscales, including social withdrawal and stereotypic behaviors did not differ significantly across children in special and mainstream settings. This further supports the conclusion that core autism symptoms (as reported by parent questionnaire rather than clinical observation) have little influence on placement decisions for autistic preschoolers.

Note that the irritability subscale of the ABC, seems to be a more clearly defined construct with strong internal consistency and conceptual clarity (Stoddard et al., 2019) that can be quantified more accurately by parents of autistic children (Kaat et al., 2014) than the other ABC subscales. Hyperactivity ABC scores, while significantly different across settings in the exploratory univariate analyses, did not reach significance in the multi-variate classification analysis. It is possible that, compared to other measures, the irritability measure more strongly reflects the children’s difficulty in regulating themselves and their reactions to the environment, which makes it difficult for them to integrate into a mainstream educational setting. In addition, impulsive reactions or angry behaviors may make it more difficult for the educational staff to provide these children with the necessary support in mainstream settings.

Finally, while some studies of school-aged children have reported that autistic children with poor adaptive abilities were more likely to be placed in special education (Towle et al., 2018) others did not (Christiansen et al., 2021; Rattaz et al., 2020; White et al., 2007). This is somewhat surprising given that children with poor adaptive behaviors including toileting, dressing, and feeding abilities are likely to require considerably more support. Nevertheless, our findings revealed that adaptive behaviors (ABAS scores) did not differ significantly across children in the two educational settings and were not informative in the classification analysis.

### Potential differences across countries

The findings of the current study may be specific to the Israeli context, where most autistic preschoolers are placed in special education settings, and approximately half receive intensive intervention programs of up to 14 weekly hours provided by a multidisciplinary team (Ilan et al., 2021; Israel Ministry of Health, 2014). In contrast, countries like the U.K. and Sweden emphasize inclusive education as the default. In the U.K., children with special needs are typically placed in mainstream settings with individualized education plans (Arnold et al., 2021), while in Sweden, nearly all preschoolers with disabilities attend mainstream programs supported by health and municipal services (Lindqvist, 2013; Sansour & Bernhard, 2018). The U.S. and Australia offer both mainstream and special education placements, but practices vary widely across regions (Altman & Barnartt, 2014; Soto-Chodiman et al., 2012). Across countries, parents are those who make the final placement decisions for their autistic children, yet a variety of differences in the availability of services and access to services are likely to impact parent decisions in different ways.

### Preschool placement policies

The results of our current and previous study (Ilan et al., 2021) highlight the lack of clearly defined evidence-based placement policies. It is not clear why cognitive function and irritability are the two main predicators of preschool educational placement. Do children with poorer cognitive abilities and higher irritability benefit more from placement in special education? Or is this an outcome of the education system’s preference to exclude children with these characteristics from mainstream education? Note that despite the significant differences in cognitive abilities and irritability across educational settings, there is still considerable overlap in these characteristics across settings such that classification accuracy is only ∼76%. In other words, there are plenty of autistic children with low cognitive scores and high irritability in mainstream education and vice versa.

This heterogeneity across educational settings will enable future longitudinal studies to examine whether children with different characteristics benefit more in one setting versus the other. Indeed, in one recent study we demonstrated that longitudinal changes in ADOS CSS over a 1-2 year period did not differ significantly across children placed in either educational setting (Ilan et al., 2021). This may seem surprising given the tremendous investment of resources that is placed in special education settings that deliver intensive autism interventions. Additional large scale longitudinal studies that compare the development of children across the two educational settings are highly warranted for determining which educational setting may benefit children with distinct characteristics, abilities, and difficulties.

### Limitations

This study has several limitations that should be addressed in future research. First, the sample was drawn from a specific geographic region in Israel, which may limit the generalizability of the findings to other cultural and geographic contexts. Comparative studies across different countries and educational systems are needed to better understand the universal and context-specific factors influencing placement decisions (Van Kessel et al., 2019). Second, the study relied on parent-reported measures of aberrant and adaptive behaviors, which may be subject to bias. Future research should incorporate multi-informant assessments, including teacher and clinician ratings, to provide a more comprehensive picture of children’s behaviors. Finally, larger nation-wide samples are needed to draw conclusive findings about factors that impact preschool placement decisions. Such an effort would require broad collaboration between academia and government offices, which is imperative for optimizing the tremendous resources that are currently being devoted to special education systems worldwide.

### Conclusions

In summary, our findings extend previous research by demonstrating that cognitive abilities and irritability are the main predictors of initial educational placement decisions for 3-year-old autistic preschoolers. At the same time, the results underscore the considerable overlap in behavioral and developmental profiles across educational settings, suggesting that placement decisions are not determined by predefined evidence-based policies, but rather by a mixture of parent intuitions and educational system preferences. Large scale, longitudinal research is desperately needed to evaluate how placement decisions impact the developmental outcomes and well-being of autistic preschoolers with different characteristics, abilities, and difficulties.

## Data Availability

All data produced in the present study are available upon reasonable request to the authors

## Notes

### Competing Interest Statement

The authors have declared no competing interest.

### Funding Statement

This study was funded by the National Autism Knowledge Center grant from the Israeli Ministry of Science and Technology.

### Author Declarations

The Soroka University Medical Center (SUMC) Helsinki Committee approved this study.

